# Cost-Effectiveness Analysis of HPV Screening Strategies in Tunisia: A modeling study

**DOI:** 10.1101/2024.10.14.24315454

**Authors:** Anas Lahdhiri, Beya Benzina, Emna Ennaifer, Ahlem Gzara, Soumaya Rammeh-Rommani, Oumaima Laraj, Hager Arfaoui, Robyn Stuart, Amira Kebir, Slimane BenMiled

## Abstract

**Background:** In Tunisia, cervical cancer ranks as the 14th most common cancer, largely driven by high-risk of Human Papillomavirus (HPV) types, notably HPV 16 and 18. Current screening efforts are limited, with only 17% of all women aged 35-60 undergoing Pap-smear testing. The introduction of the HPV vaccine in 2025 through the national school vaccination program, targeting girls aged 11-12, is expected to reduce the burden of cervical cancer. However, alongside vaccination, enhanced screening strategies are essential for early detection and prevention of HPV-related cancers.

**Aim:** This study aims to assess the cost-effectiveness of different HPV screening strategies in Tunisia, specifically examining the combination of varying HPV screening frequencies and a vaccination program targeting girls aged 11-12. The objective is to determine the most cost-efficient screening strategy to complement vaccination efforts in reducing cervical cancer incidence.

**Methods:** A cost-effectiveness analysis was conducted from the perspective of the Tunisian healthcare system using the HPVsim model, a multi-agent-based simulation tool that captures HPV transmission dynamics and cervical cancer progression. Four approaches were compared: (1) maintaining the current Pap-smear screening strategy combined with vaccination; (2) introducing HPV DNA testing once between ages 35-40; (3) introducing HPV DNA testing twice between ages 35-45, with a 5-year interval; and (4) introducing HPV DNA testing every 5 years for women aged 35-60. All approaches were combined with the vaccination program. Screening coverage rates of 15%, 25%, 33%, 50%, and 70% were tested for each approach. Primary outcomes included the number of cancer cases averted, total intervention costs, and cost increase per cancer case averted. Academic literature and existing evidence were included on the demographic variables, cervical cancer incidence and mortality, treatment costs, vaccine delivery costs and other model parameters.

**Results:** All interventions resulted in substantial reductions in cervical cancer cases, with decreases ranging from 41% to 59% in cumulative cases between 2025 and 2090. The most intensive approach, involving HPV DNA testing every 5 years for women aged 35-60, achieved the largest reduction, with a 59% decrease in cumulative cervical cancer cases by 2090, although it also incurred the highest costs. The least costly option, which retained current Pap-smear testing alongside vaccination, reduced cervical cancer cases by 41%. Although the introduction of HPV DNA testing significantly increases costs, a high frequency of screening allows for quicker public health benefits. The scenario combining vaccination and maintaining current screening practices is found to be the most cost-effective for the Tunisian context. If the price of the HPV DNA test is reduced to $9 USD, the most frequent testing strategy would become the most cost-effective option, offering both high effectiveness and financial viability.

**Conclusion:** Lowering the cost of HPV DNA testing could make more frequent screening financially sustainable, providing greater public health benefits. These findings offer valuable guidance for decision-makers in shaping future strategies for cervical cancer prevention in Tunisia.

## Introduction

Human Papillomavirus (HPV) is a group of viruses transmitted through sexual contact, infecting the epithelial cells of the skin and mucous membranes, HPV infections can lead to benign conditions like genital warts and, over time, to precancerous or cancerous lesions, particularly uterine cervical cancer^1^.

In Africa, despite ongoing efforts to enhance prevention and treatment, cervical cancer continues to be a major concern due to socioeconomic challenges, restricted healthcare access, and insufficient awareness^1^. To address this, the World Health

Organization (WHO) has launched the “90-70-90” initiative, which aims to globally eliminate cervical cancer by vaccinating 90% of girls against HPV before age 15, screening 70% of women at ages 35 and 45, and treating 90% of women with cervical disease^2^. This strategy is supported by awareness campaigns, international collaborations, and funding to bolster health systems in developing countries.

The WHO advocates for HPV screening as a key tool for early detection of cervical cancer, particularly in low- and middle-income countries. The guidelines recommend that women start screening at age 30 and continue at regular intervals, typically every 5 to 10 years, depending on the type of test and previous results^2^. In Tunisia, the Pap smear test is used for only 1% of women aged 35 to 60 years, largely due to the limited capacity of laboratories. As a result, there is growing interest in adopting the HPV DNA test, which offers automation and ease of processing. Although this test primarily detects HPV types 16 and 18, it could significantly increase screening coverage. HPV DNA test prices vary by manufacturer (Hologic Aptima, GenProbe Aptima, BD Onclarity, CareHPV) and region (low-income coutries, middle-income countries) from $5 to $100 ^3 4 5^. Given the high cost of testing, it is essential to evaluate screening frequency and coverage alongside vaccination efforts.

In Tunisia, HPV types 16 and 18 are the most common genotypes linked to cervical infections and cancers, with other high-risk types such as HPV 31, 45, 51, and 56 also prevalent^3^. Cervical cancer is the 14^*th*^ most common cancer, representing 2% of cancers in Tunisia in 2022^4^. Starting in 2025, vaccination will be incorporated into the existing school program, using the bivalent vaccine to target girls aged 11 to 12, with an estimated coverage rate of 90% ^6^. A cost-effectiveness analysis has been conducted to compare various vaccination strategies which demonstrate that CECOLIN vaccine emerges as the most cost-effective HPV vaccine, particularly when cross-protection is considered -the willingness-to-pay (WTP) threshold of 5% of GDP^5^.

The goal of this work is to identify the most cost-efficient screening strategy to complement vaccination efforts. We will use the multi-agent-based model HPVsim, which integrates social interaction dynamics, HPV transmission, and its progression to cervical cancer.

The document presents a cost-effectiveness analysis of combined HPV vaccination and screening strategies in Tunisia. The section details the use of the HPVsim model for simulation, including descriptions of intervention scenarios, costs, and outcomes. It covers demographic, biological, and social parameters, focusing on varying screening frequencies and vaccine coverage. The section compares the effectiveness of different strategies in reducing HPV infections and cervical cancer cases. Finally, section concludes the document.

## Results

We simulated the maintenance of the current cervical cancer screening process while simultaneously introducing routine vaccination for girls aged 11–12 (see Figure 1b and 1a in red). The simulation projected a reduction in both HPV infections and cervical cancer cases by 65% and 76%, respectively, by the year 2090. We also evaluated alternative strategies defined by scenarios 2, 3 and 4. By 2090, Scenario 2, which introduced HPV DNA testing at a frequency of one test for women aged 35–40 plus routine vaccination, showed reductions in HPV infections and cervical cancer cases by 65-67% and 75-77%, respectively (see Figures 1b and 1a in blue). Scenario 3, which introduced HPV DNA testing at a frequency of two tests for women aged 35–45 plus routine vaccination, showed reductions in HPV infections and cervical cancer cases by 62-68% and 74-79%, respectively (see Figures 1b and 1a in yellow). Finally, Scenario 4, which introduced HPV DNA testing at a frequency of every 5 years for women aged 35–60 plus routine vaccination, showed reductions in HPV infections and cervical cancer cases by 63-66% and 75-79%, respectively (see Figures 1b and 1a in green). The slight upward trend observed at the beginning of the curves for scenarios 1 and 2 in Figure 1b (red and blue, respectively) is due to the convex shape of the Baseline curve, which shows a continuous and accelerating increase in yearly cancer cases. In contrast, the scenario curves are concave, initially increasing but then declining due to vaccination and/or screening interventions. As the model is stochastic, smoothing them against the Baseline causes this effect. The minor upward peak, under 5%, is negligible.

**Figure 1.**
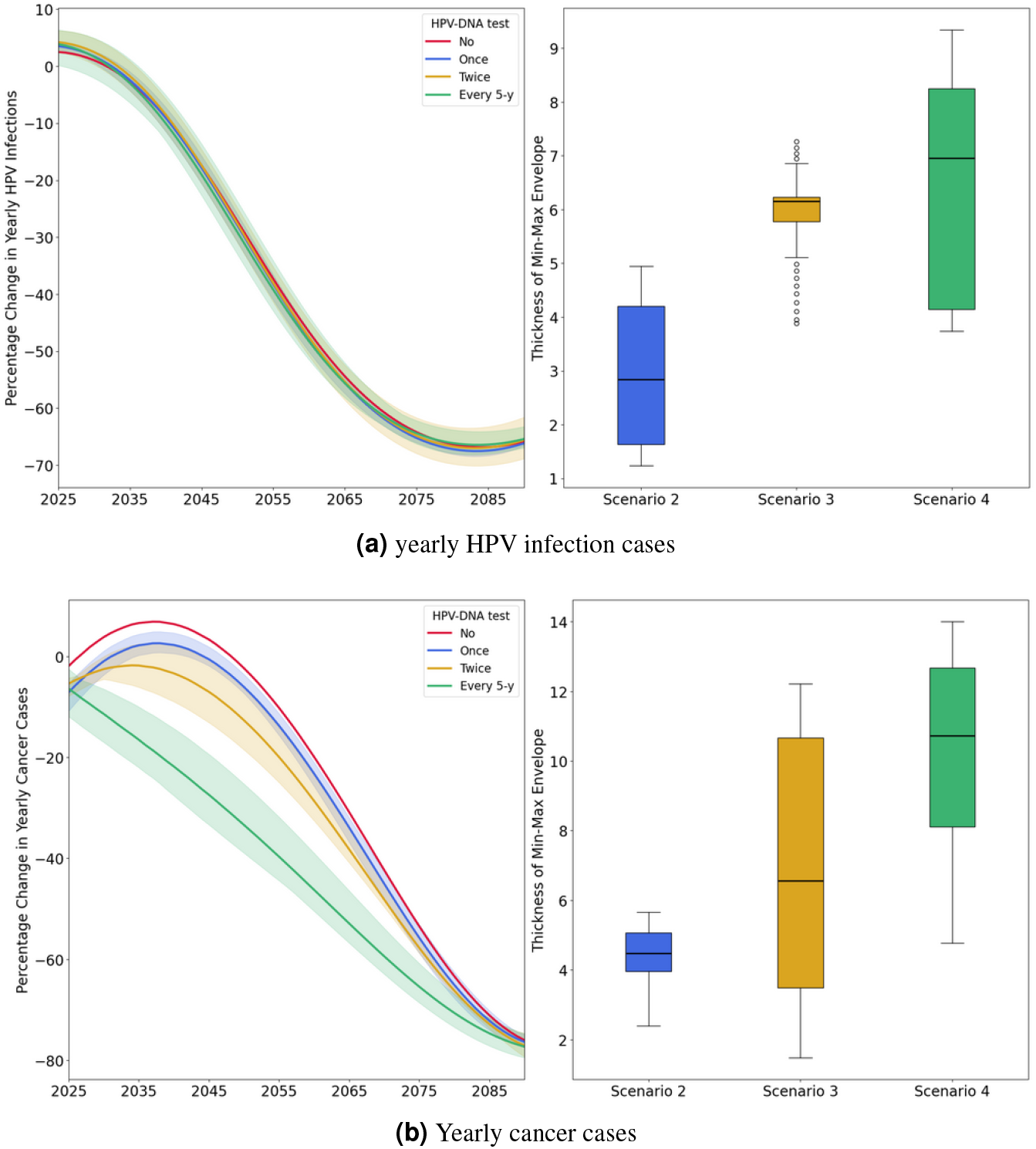
The annual reduction in HPV infections and cancer cases for scenarios (1-4), compared to the baseline scenario. For scenarios (2-4), the curves represent the average reductions across various coverage rates tested, with the shaded areas indicating the range of minimum and maximum values (min-max envelope) resulting from different coverage rates. The boxplots display the thickness of the min-max envelope, reflecting the variability due to the different coverage rates tested for each strategy. Scenario 1 (red): maintaining the current screening strategy coupled with vaccination; Scenario 2 (blue): introducing the HPV DNA test at a frequency of one screening between ages 35 and 40, coupled with vaccination; Scenario 3 (yellow): introducing the HPV DNA test at a frequency of two screenings spaced 5 years apart between ages 35 and 45, coupled with vaccination; Scenario 4 (green): introducing the HPV DNA test at a frequency of every 5 years for women aged 35-60, coupled with vaccination.

Although all scenarios resulted in almost similar reductions in HPV infections and cervical cancer cases by 2090, the rate of decrease differed across scenarios. The more frequent is the screening, the steeper the decline in the annual cancer cases curve. For instance, the annual cancer cases curve for Scenario 1 begins its downward trend from 2040, for Scenario 2 from 2040, for Scenario 3 from 2035, and for Scenario 4 almost immediately from 2025 (see Figure 1b). However we don’t notice any considerable difference in the rate of decrease of HPV infections between all scenarios. For further examination, supplementary analysis was conducted to study the effect of HPV-DNA test introduction by comparing the reduction in HPV infections and cervical cancer cases of scenarios 2-4 which include the HPV DNA test to the scenario 1 which doesn’t include the introduction of HPV DNA test. For reduction in HPV infections there are no notable differences between scenarios 2-4 as all the curves are within a 0%-7% difference margin (see Figure 2a). As for reduction in cancer cases there are notable differences as the more frequent the screening, the more we notice the effect of the introduction of HPV-DNA test; for instance in scenario 2 where the HPV-DNA test is conducted at a rate of once in a lifetime there is barely any noticeable effect when compared to scenario 1 which doesn’t include the introduction of HPV-DNA test as reduction of cancer cases of scenario 2 compared to scenario 1 is always nearing 0% (see Figure 2b), in scenario 3 where the HPV DNA test is conducted at a rate of twice in a lifetime, the reduction of cancer cases when compared to scenario 1 reaches a maximum of 11%, and in scenario 4 where the HPV DNA test is conducted at a rate of every 5 years, the reduction of cancer cases when compared to scenario 1 reaches a maximum of 33%. We observe that over longer time horizons, particularly by the year 2090, the outcomes will be primarily influenced by vaccination rates, as the impact of introducing the HPV-DNA test diminishes, and by 2090, all scenarios become equivalent.Despite the fact that the more frequent is the screening, the more noticeable the effect of the introduction of HPV-DNA test, we see that at the long run this effect is reduced and by the year 2090 all scenarios are equivalent.

**Figure 2.**
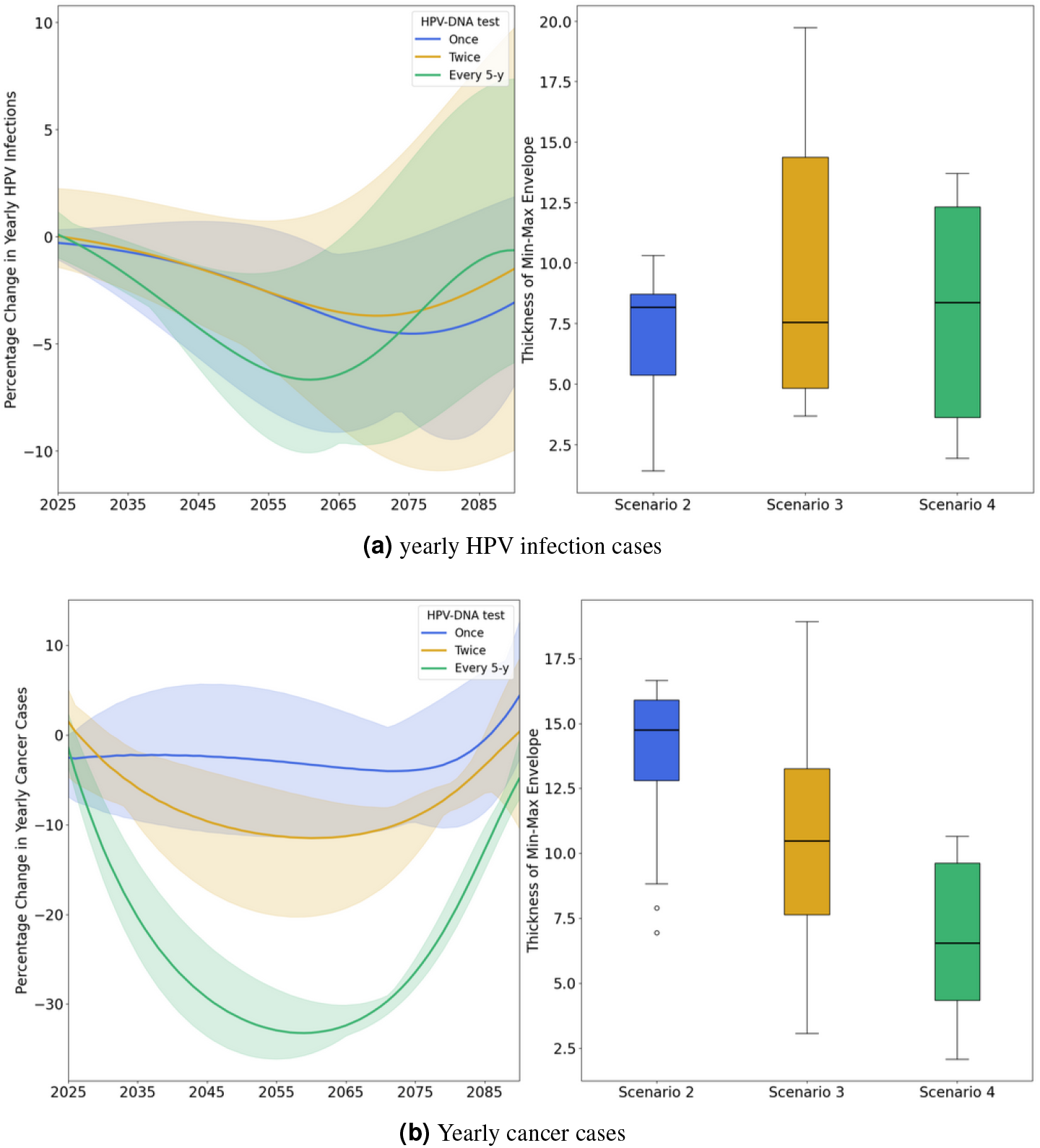
The annual reduction in HPV infections and cancer cases for scenarios (1-4), compared to the scenario 1. For scenarios (2-4), the curves represent the average reductions across various coverage rates tested, with the shaded areas indicating the range of minimum and maximum values (min-max envelope) resulting from different coverage rates. The boxplots display the thickness of the min-max envelope, reflecting the variability due to the different coverage rates tested for each strategy. Scenario 2 (blue): introducing the HPV DNA test at a frequency of one screening between ages 35 and 40, coupled with vaccination; Scenario 3 (yellow): introducing the HPV DNA test at a frequency of two screenings spaced 5 years apart between ages 35 and 45, coupled with vaccination; Scenario 4 (green): introducing the HPV DNA test at a frequency of every 5 years for women aged 35-60, coupled with vaccination.

In a second analysis, we examined the percentage of cumulative cancer cases averted from 2025 to 2090 across these different scenarios. Results indicate that the more frequent is the screening, the higher the percentage of cumulative cancer cases averted. We should note that varying the screening coverage rate within the same scenario had little influence on the outcome in terms of cancer reduction, as there was no significant difference in the mean percentage of cumulative cancer cases averted (see Figure 3). Scenario 1 resulted in a reduction of cumulative cancer cases from 2022 to 2090 by 41%. In scenario 2 the mean percentage of cumulative cancer reduction varies between 42% and 47%. In Scenario 3 the mean percentage of cumulative cancer reduction varies between 44% and 49%. Finally, in Scenario 4 the mean percentage of cumulative cancer reduction varies between 52% and 59% (see Figure 3).

**Figure 3.**
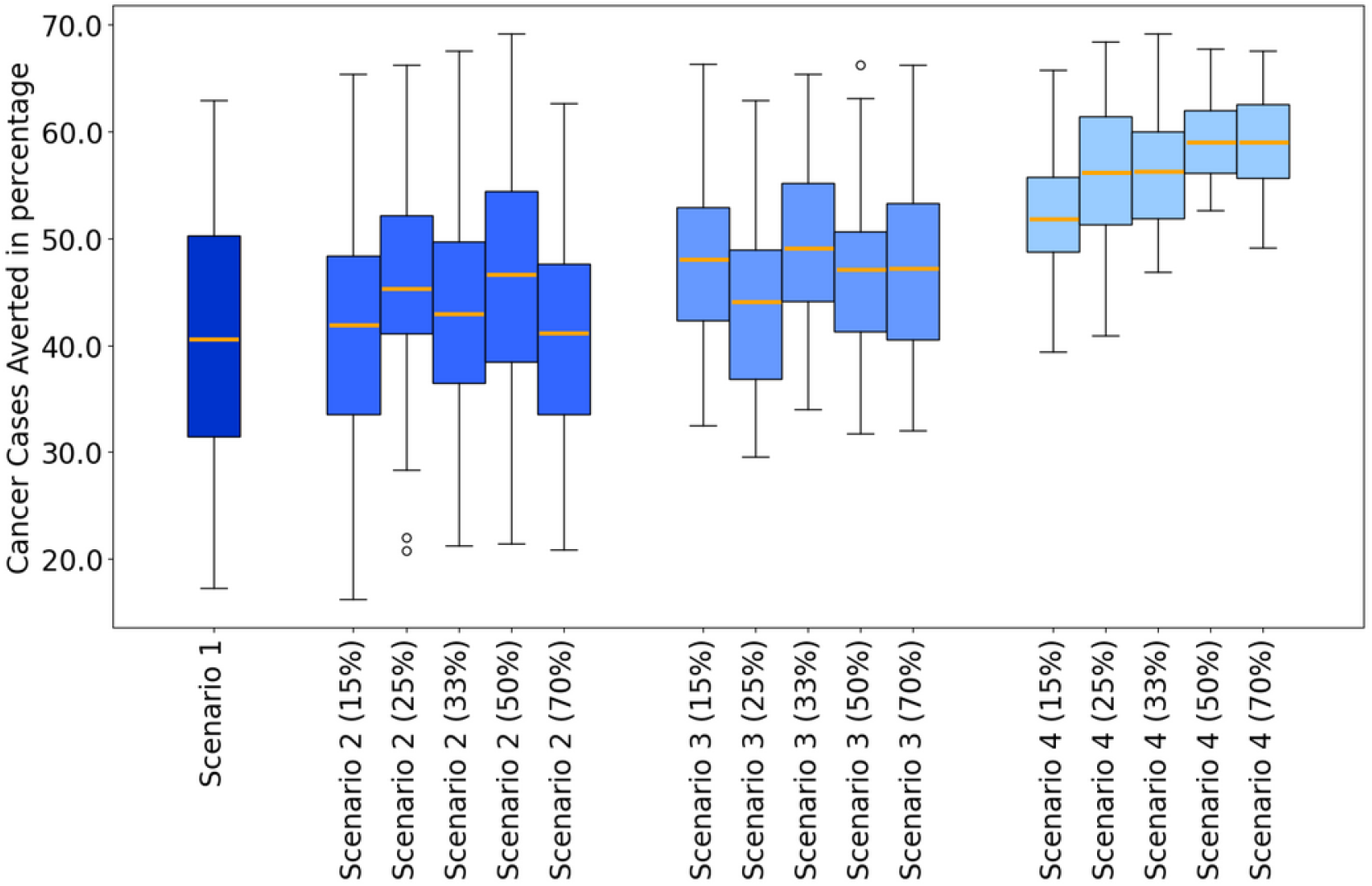
The figure presents the cumulative cancer cases averted percentage for each scenario compared to the baseline scenario. Error bars are the 90th and 10th percentile, boxes are the 75th and 25th percentiles, horizontal lines in each box is the mean. Scenario 1 is maintaining the current screening strategy coupled with vaccination; Scenario 2 is introducing the HPV DNA test at a frequency of one screening during a lifetime at the age between 35 and 40, coupled with vaccination; Scenario 3 is introducing the HPV DNA test at a frequency of two screenings during a lifetime spaced 5 years apart at the age between 35 and 45, coupled with vaccination; Scenario 4 is introducing HPV DNA test at a frequency of every 5 years for women aged between 35 and 60, coupled with vaccination.

Cost analysis reveals that scenarios incorporating HPV DNA testing are significantly more expensive compared to Scenario 1, which retains the less-costly Pap-smear test. Figure 4 shows that costs escalate with greater coverage rates within each scenario which is contrary to what we noted on the effect of increasing coverage rate on reducing cancer cases. We also notice that Scenarios 1 and 2 are cost-saving, suggesting that these two scenarios are less expensive than the Baseline Scenario therefore adapting one of these two strategies will actually generate positive revenue.

**Figure 4.**
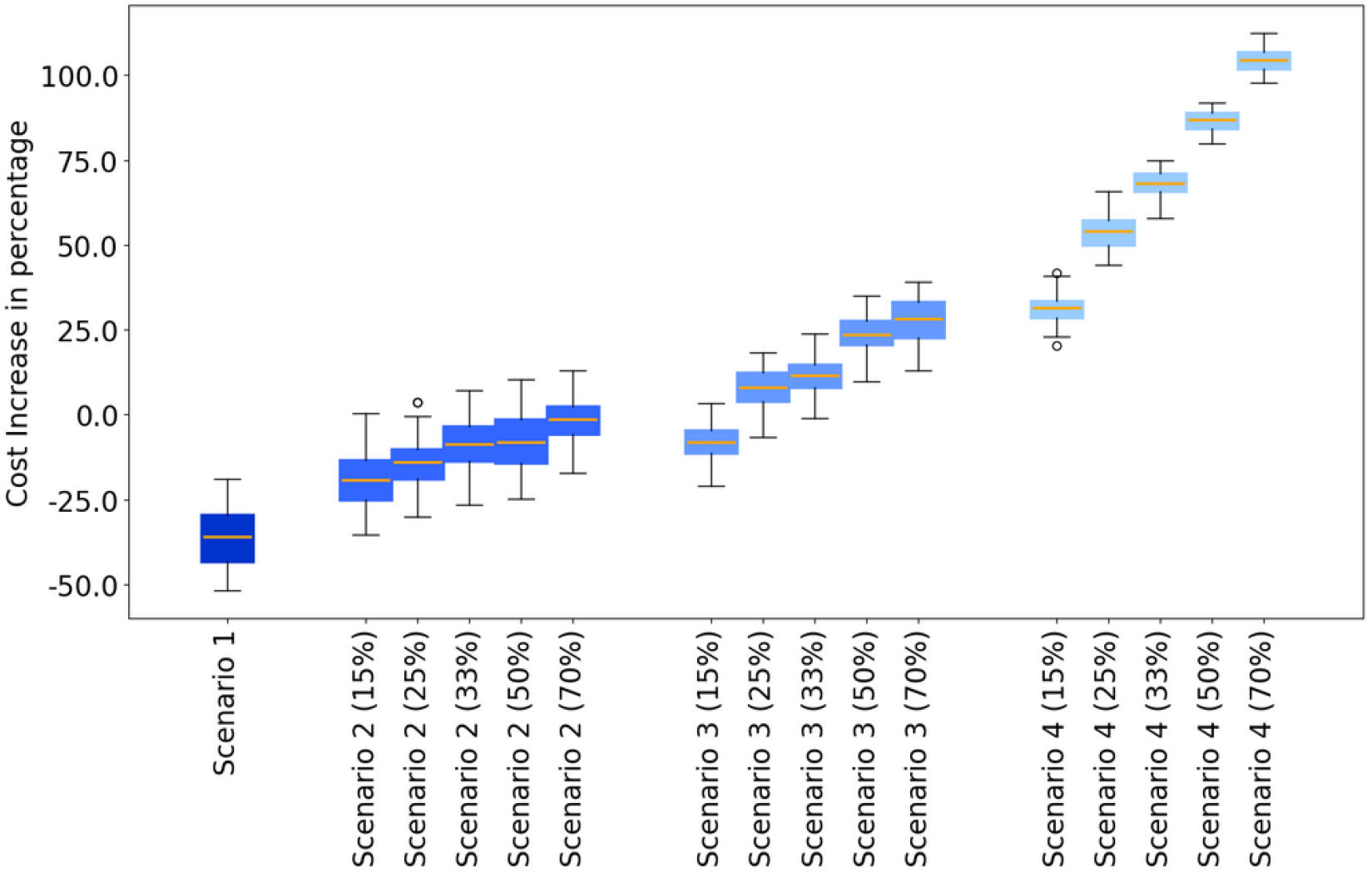
The figure presents the cost increase in percentage for each scenario compared to the baseline scenario. Error bars are the 90th and 10th percentile, boxes are the 75th and 25th percentiles, horizontal line in each box is the mean. Scenario 1 is maintaining the current screening strategy coupled with vaccination; Scenario 2 is introducing the HPV DNA test at a frequency of one screening during a lifetime at the age between 35 and 40, coupled with vaccination; Scenario 3 is introducing the HPV DNA test at a frequency of two screenings during a lifetime spaced 5 years apart at the age between 35 and 45, coupled with vaccination; Scenario 4 is introducing HPV DNA test at a frequency of every 5 years for women aged between 35 and 60, coupled with vaccination.

Combining cost and efficacy data, we compared the cost increase per cancer case averted for each scenario to the average GDP between 2025 and 2090. The cost-effectiveness analysis reveals that all strategies 1-4 falls within the “cost-effective” range, (see Figure 5a) as all scores are under the three times the average GDP per capita from 2025 to 2090 threshold. Furthermore, strategies 1-2 have negative cost-effectiveness scores meaning that adapting one of these strategies is cheaper than having no intervention at all. Scenario 3 is emerging as “very cost effective” because its scores are between zero and one times the average GDP per capita. Finally Scenario 4 is emerging as “cost effective” because its scores are between one times and three times the average GPD per capita.

**Figure 5.**
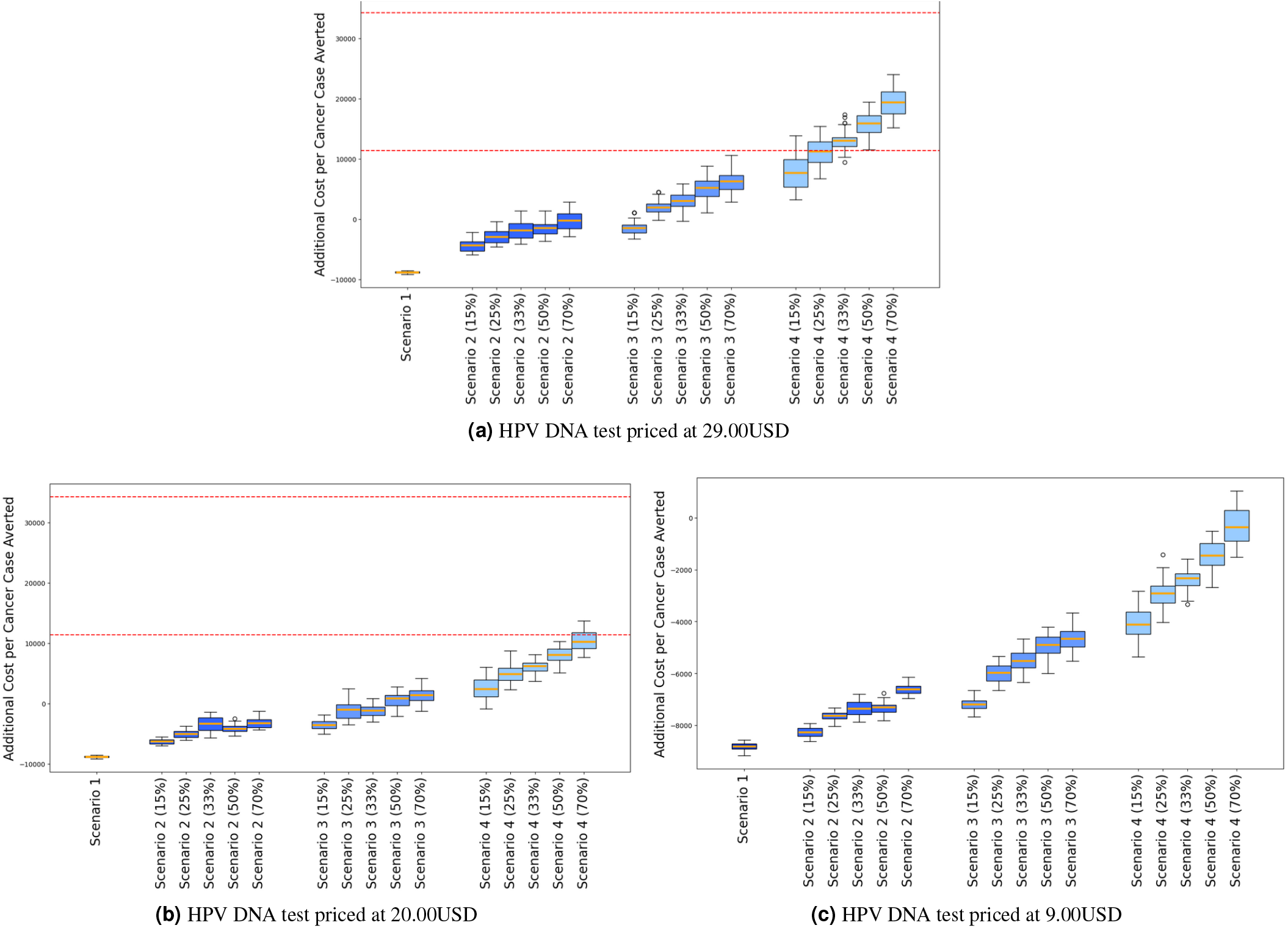
The figure illustrates the cost-effectiveness defined as the cost increase per cancer case averted for all the screening strategies across all the tested coverage rates of screening at different prices of the HPV DNA test. Error bars are the 90th and 10th percentile, boxes are the 75th and 25th percentiles, and horizontal lines in each box represent the mean. The dotted horizontal lines represent the “cost-effective” range defined as a score between one and three times the average GDP per capita between 2025 and 2090. Scenario 1 is maintaining the current screening strategy coupled with vaccination; Scenario 2 is introducing the HPV DNA test at a frequency of one screening during a lifetime at the age between 35 and 40, coupled with vaccination; Scenario 3 is introducing the HPV DNA test at a frequency of two screenings during a lifetime spaced 5 years apart at the age between 35 and 45, coupled with vaccination; Scenario 4 is introducing HPV DNA test at a frequency of every 5 years for women aged between 35 and 60, coupled with vaccination.

**Figure 6.**
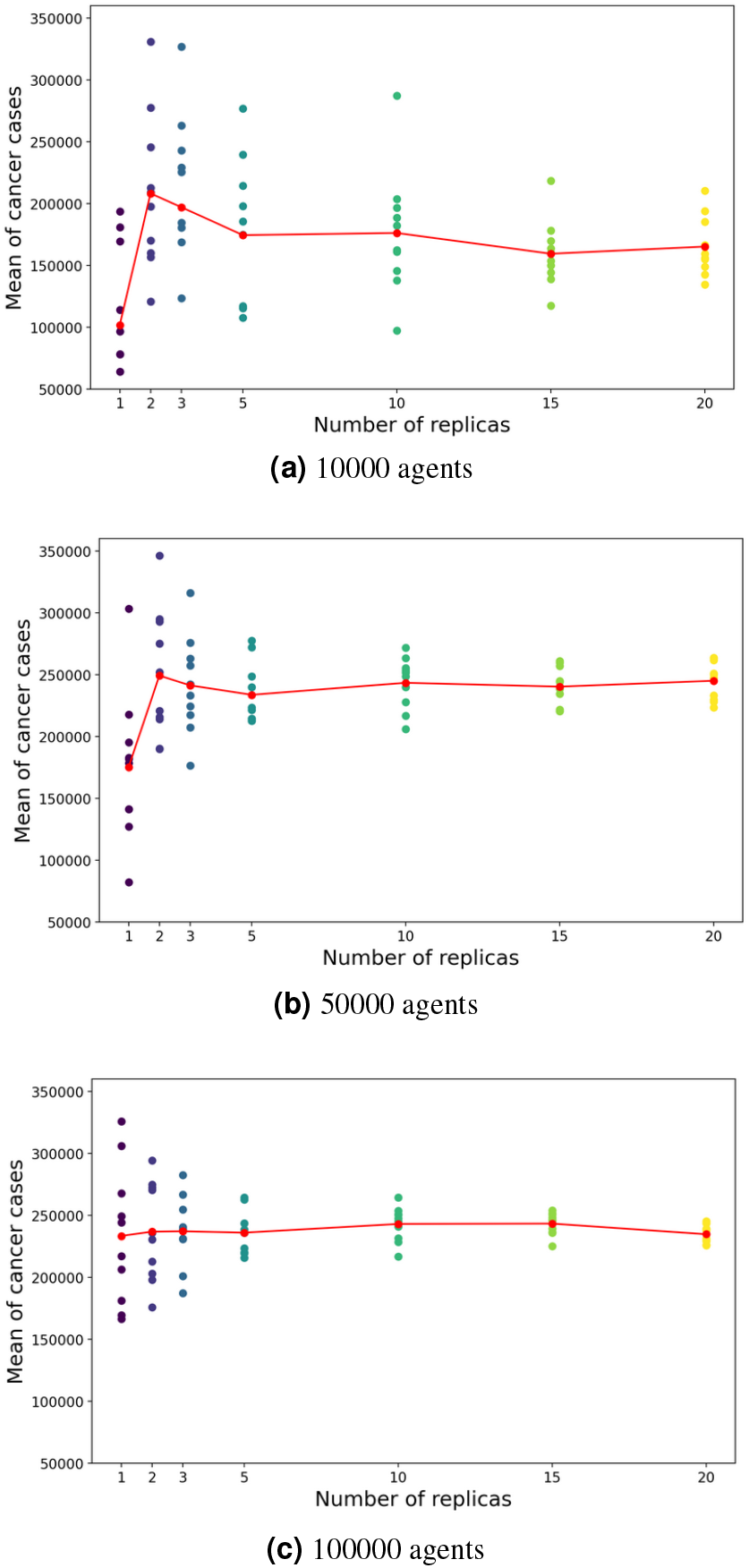
The figure presents scatter plots representing the mean cancer cases for each number of replicates across the different agent population sizes. The red curve traces the centroid of each scatter plot (i.e., mean of means).

**Figure 7.**
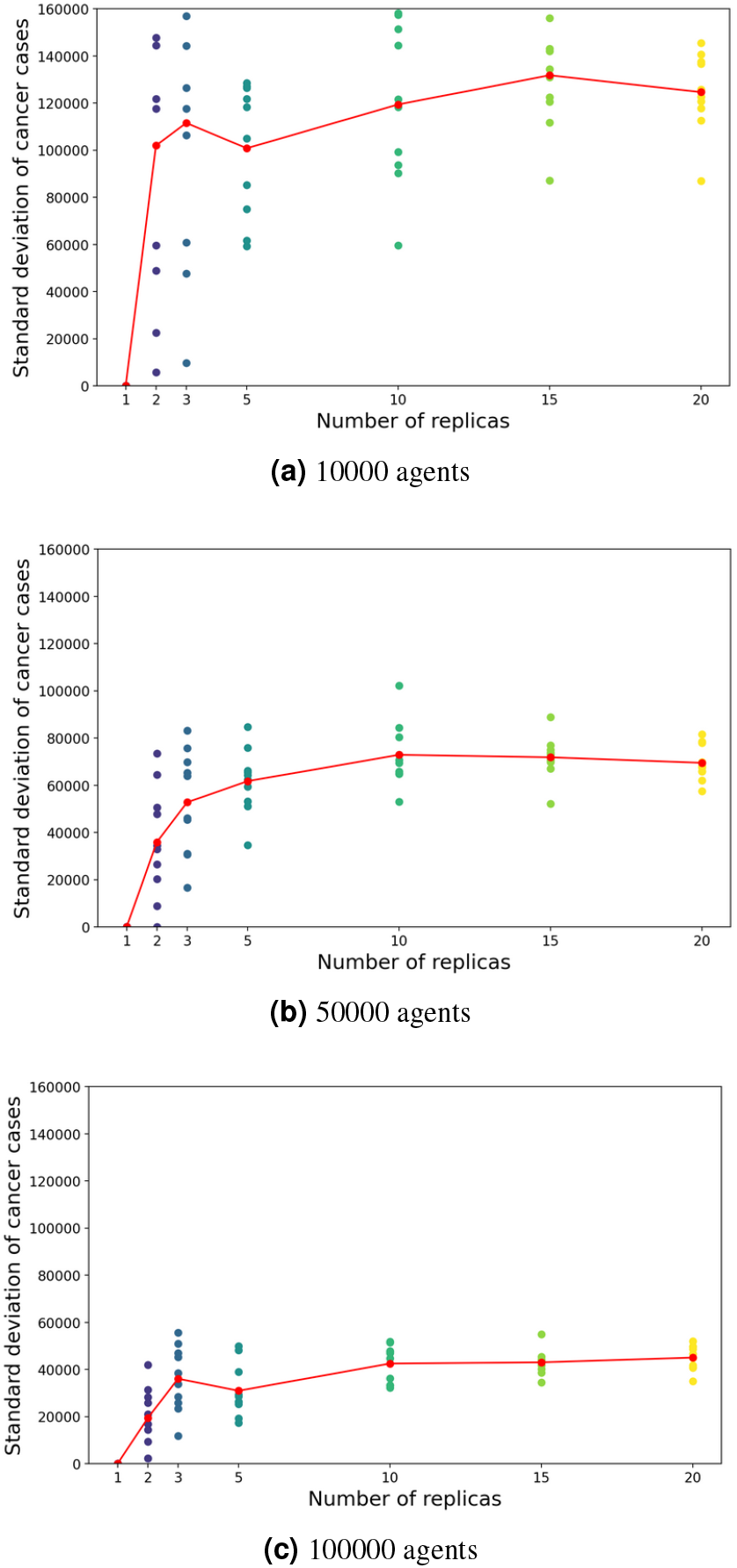
The figure presents scatter plots representing the standard deviation of cancer cases of each number of replicates across the different agent population sizes. The red curve traces the centroid of each scatter plot (i.e., mean of standard deviations).

**Figure 8.**
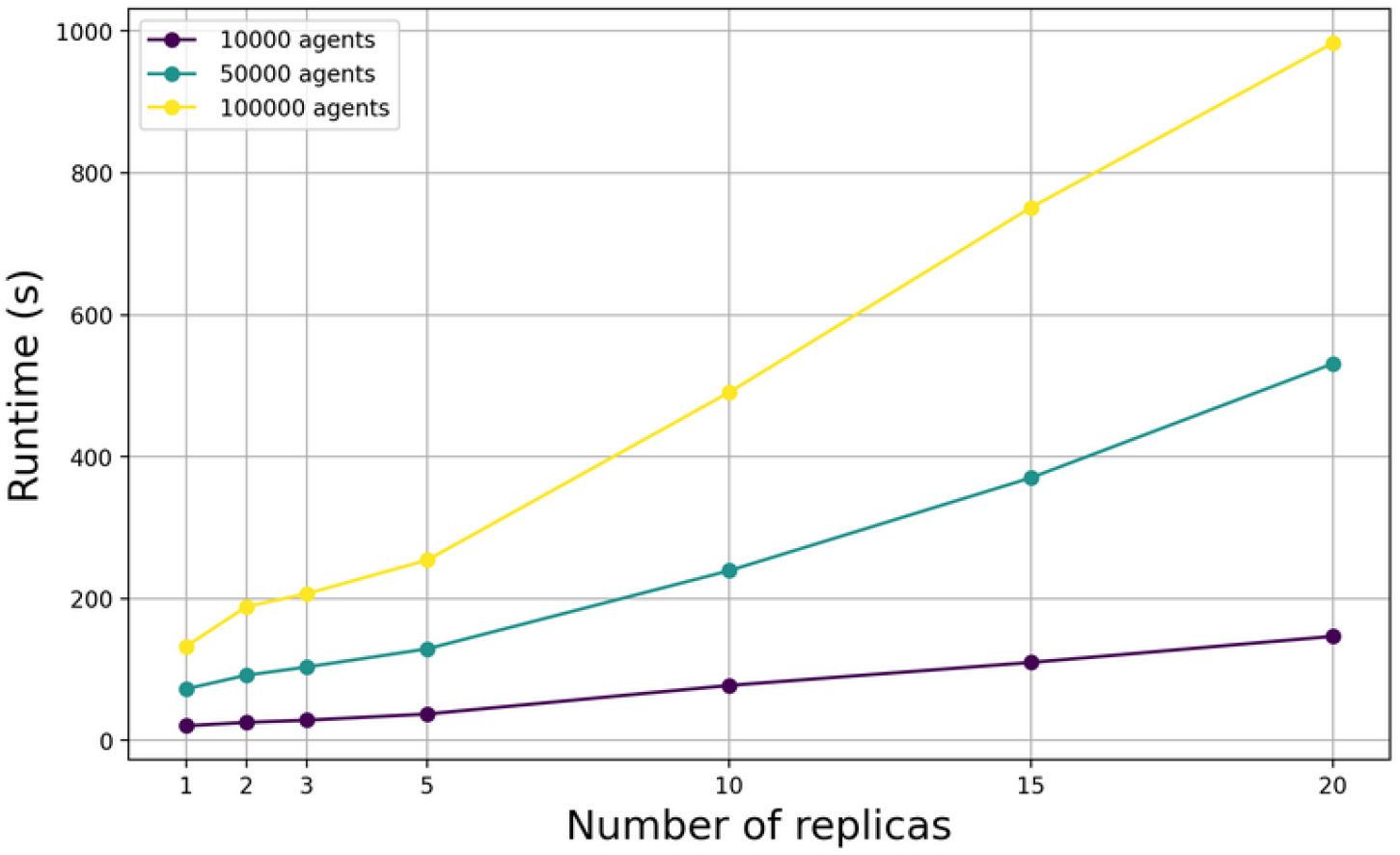
Running time of simulations for different agents populations sizes as a function of the number of replicates to perform

**Figure 9.**
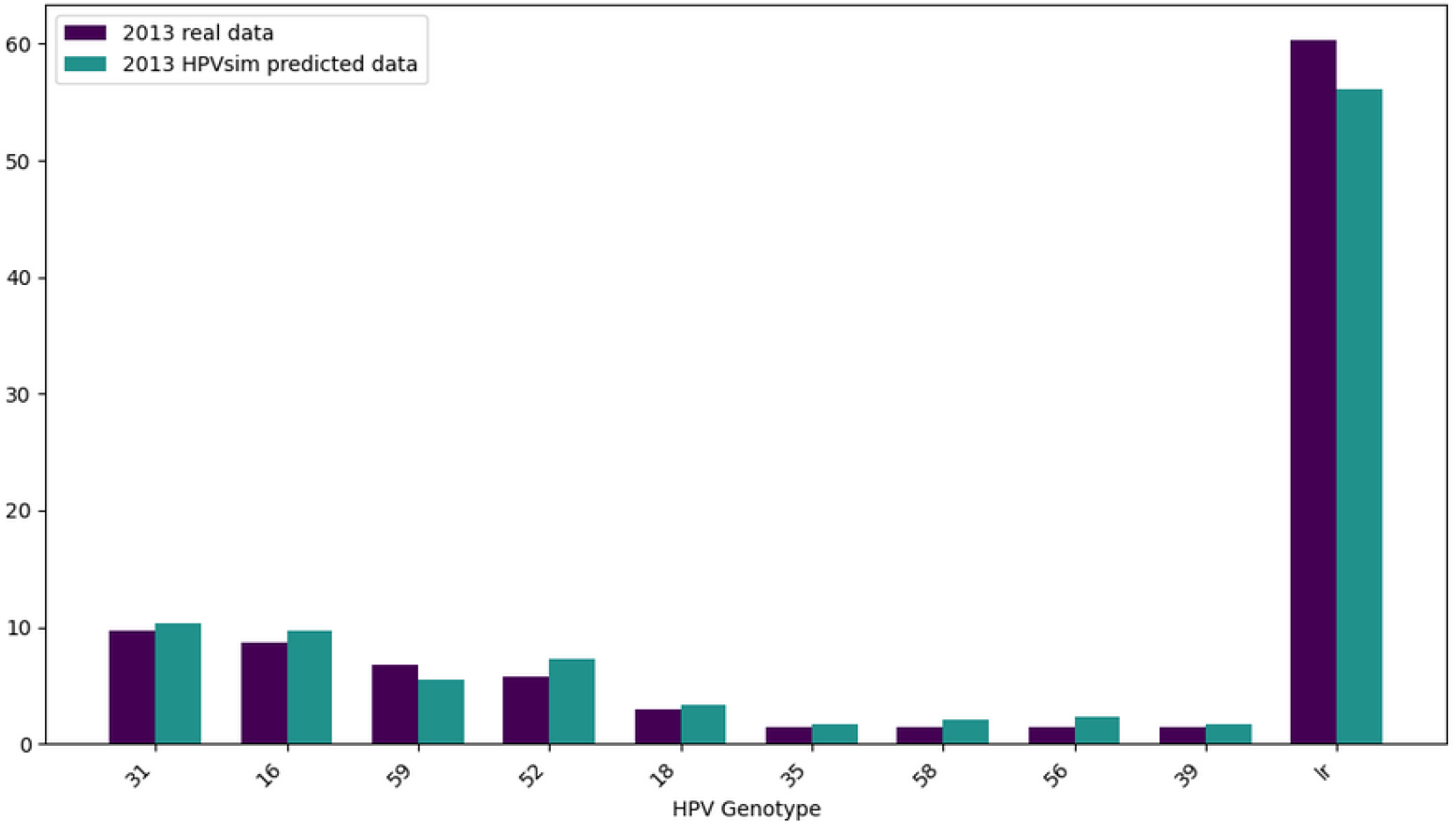
Barplot of genotype prevalence in the population in 2013, in violet the distribution according to the real data and in green the distribution resulted from the simulation

**Figure 10.**
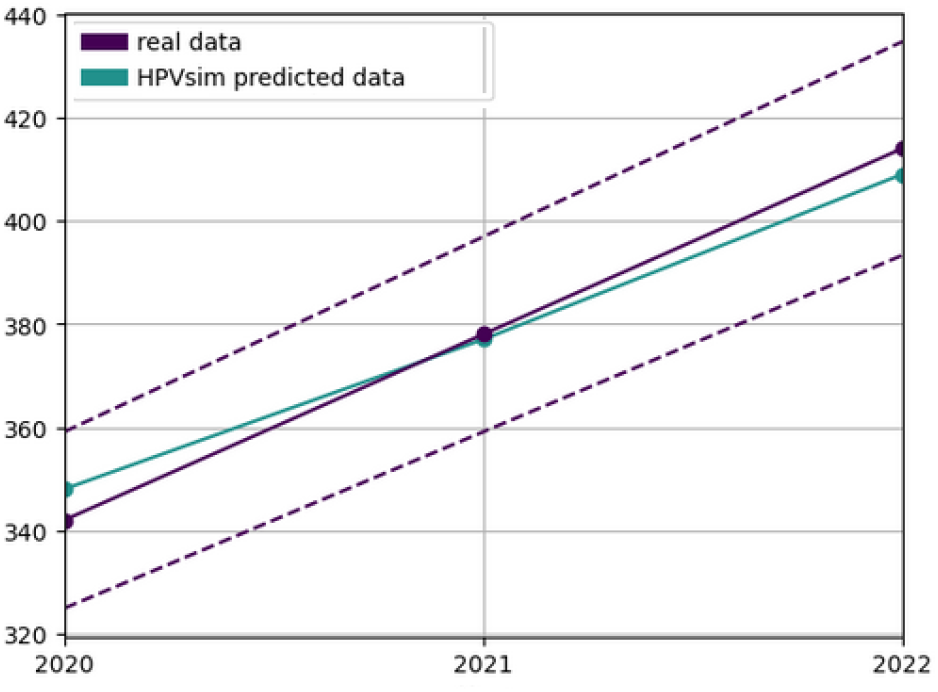
The figure represent the yearly cancer cases from 2020 to 2022, in violet the real data, in green the predicted yearly cancer cases by HPVsim and the dotted lines represent the 10% error margin

A sensitivity analysis was conducted to explore the feasibility of strategy 4 that includes screening at a rate of every 5 years for women aged between 35 and 60. Our findings suggest that at a price of 20.00USD for the HPV-DNA test, scenarios 1-3 are cost-saving while Scenario 4 is henceforth in the “very cost effective” range (see Figure 5b). Furthermore, if the price of the HPV DNA test could be reduced to 9.00 USD or less, all scenarios are cost-saving whatever the coverage rate is (see Figure 5c), this suggests that adapting any of the scenarios is cheaper than the Baseline scenario. In this case, Scenario 4, which includes vaccination for girls aged 11–12 and HPV DNA screening every 5 years for women aged 35–60, specifically at a coverage rate of 70%, emerging as the most favorable due to its highest reduction (see Figure 3) in cancer case.

## Discussion

The association between cervical cancer and the HPV genome was first identified in 1983, marking a turning point in cervical cancer research^6^. Since then, significant progress has been made in the fight against the disease. However, after the HPV vaccine was approved for clinical use in June 2006, the focus of the scientific and medical community shifted from the simple prevention of cervical cancer to the ambitious goal of eliminating it^7^.

Our current study demonstrated that the most cost-effective strategy for the Tunisian context towards that goal, involves introducing routine HPV vaccination with the bivalent vaccine for girls aged 11 to 12 years, achieving a 90% coverage rate, while maintaining the current screening strategy using Pap-smear tests for women aged 35 to 65 years with a 1% coverage rate. This approach is projected to reduce cumulative cervical cancer cases by 41% between 2025 and 2090. By 2090, the reduction in cancer cases is expected to reach 76% compared to the baseline scenario without vaccination. Although strategies that include HPV DNA screening method coupled with vaccination are also cost-effective. Regarding screening tools, our findings align with those of Chen M-K et al., who demonstrated that annual Pap smear testing yields the highest gain in life years when compared to HPV DNA testing^8^. This could be explained by the fact that Pap smear screening not only detects early-stage invasive cervical carcinoma but also prevents the malignant transformation of precursor lesions. Additionally, the cost of Pap smear testing is lower than that of HPV DNA testing, making it a more cost-effective option. On the other hand, the WHO recommends HPV DNA based screening, as it is less prone to human errors and also suitable for all settings, areas, and countries ^7^. Moreover, cost-effectiveness studies done in other low and middle income countries (LMIC) including India, Kenya, Peru, South Africa and Thailand confirm the efficacy of HPV DNA screening and its cost effectiveness in these areas^9^. The divergence between these findings highlights the importance of considering country-specific factors when choosing screening strategies. Differences in healthcare infrastructure, the availability of healthcare professionals, gross domestic product (GDP), and the population’s acceptance of certain screening tools all play critical roles in determining the most suitable approach for each country. In Tunisia, despite the low coverage rate due to limited capacity of labs to process tests, the geographical distribution is good across the country with 2085 primary care centers distributed throughout the 24 governorates^8^. When it comes only to the number of cancer cases, our results shows that maintaining the current screening strategy with existing coverage rates in Tunisia or introducing intensive screening based on the HPV DNA test yields similar outcomes by the end of our time horizon when compared to the introduction of the HPV DNA test. Actually, the key difference lies in the time required to achieve these results and the deployed costs. The strategy involving intensive HPV DNA screening significantly accelerates the reduction in cervical cancer cases, leading to earlier public health benefits’ range although being costlier than the first scenario. As explained by Castaneda-Orjuela et al, the decision making process in the public health field should be a comprehensive process valuing consequences beyond the health system, including the population well-being^10^.

Our study has several notable strengths. First of all, the HPVsim model was parameterized to closely align with Tunisian demographic and behavioral parameters, as well as the prevalence of HPV in the country. This customization ensures that the model accurately reflects the unique characteristics of the Tunisian population. Secondly, the model was rigorously calibrated to produce predictions that closely match real-world data, such as the distribution of HPV genotypes in 2013 and the annual incidence of cervical cancer cases between 2020 and 2022. This calibration process enhances our confidence in the relevance and reliability of the results within the Tunisian context. Lastly, all the scenarios and their variations were developed in close collaboration with experts in vaccination and HPV screening from the Pasteur Institute of Tunis. Their insights, suggestions, and feedback were invaluable in ensuring that the study’s questions were pertinent to real-world challenges. This collaboration significantly contributes to the practical applicability of our findings, aiding in shaping the decision-making process regarding HPV vaccination and screening strategies in Tunisia.

Our study also has several limitations that should be taken into account. Firstly, we did not explore all possible HPV screening tests available on the market, nor did we consider every potential screening target or routine vaccination target and coverage. This decision was made to focus on strategies that were most relevant to Tunisian policymakers at the time of the study. Secondly, we did not incorporate the effects of migration or HIV infections into our model. This omission could potentially lead to an overestimation of the vaccine’s effectiveness. Nevertheless, we think that despite these limitations, the overall conclusion of the study should remain robust as the most cost-effective strategy identified would likely remain the same, though the exact efficacy score might differ. Lastly, when calculating costs, we assumed a constant discount rate of 3% throughout the entire simulation period, extending until 2090. While this is a simplified assumption, it’s a common assumption used in many studies of cost-effectiveness analysis in public health^11^–15 as it’s recommended by the Second Panel on Cost-Effectiveness in Health and Medicine^16^.

Models can be categorized by the following criteria: (a) Randomness: Deterministic models use pre-determined rates for events such as HPV acquisition and clearance, resulting in consistent outcomes, while stochastic models incorporate randomness, leading to varying results with each run. (b) Structure Level: Individual-based models simulate events for each person, capturing individual variability, whereas aggregate models group individuals into compartments, reducing internal variability. (c) Interaction: Dynamic models consider herd immunity by modeling infection risk based on the number of infectious individuals, reflecting changes over time. In contrast, static models assume a constant infection rate and do not account for herd immunity. HPVSim is a dynamic, stochastic multi-agent model that explicitly incorporates both vaccination and screening, along with complex social sexual interactions^17^. This integration of multiple factors makes it an ideal choice for our study, providing a detailed and realistic simulation of HPV transmission dynamics and preventive measures. It is worth noting that similar models exist, such as those by^18^,19. However these models, HPVsim included, do not consider certain criteria such as equity and acceptability. In the aftermath of the COVID pandemic, vaccine hesitancy has become a significant threat to global health^20^. Experts are increasingly recommending that the acceptability of vaccines and screening tools be included in health economic evaluations, which was not done in this study^21^.

## Methods

### Study design

We conducted an economic evaluation through a cost-effectiveness analysis of combined vaccination and screening strategies tailored to the Tunisian context. This analysis was approached from a healthcare system perspective using the multi-agent model HPVsim. The analysis was reported according to the Consolidated Health Economic Evaluation Reporting Standards statement.

### Modelling

HPVSim is a software package developed that provides flexibility for creating agent-based models to simulate HPV transmission and cervical disease progression^17^. The simulations generated using HPVSim feature a population of agents who interact via sexual networks, over which multiple co-circulating HPV genotypes can be transmitted. Persistent infections can lead to cervical dysplasia, which may spontaneously regress or progress to invasive cervical cancer. HPVSim can also simulate the effects of interventions, including vaccination, screening, and treatment.

By default, simulations run with HPVsim include genotypes 16, 18, and a pooled group consisting of all other high-risk types. In Tunisia, studies^3^ has shown that many high-risk genotypes such as HPV 31, 35, 39, 52, 56, 58 and 58 are prevalent in the population. Therefore we modified to the HPVsim code to include these additional high-risk genotypes. A detailed description on genotype identification is available in appendix.

The model parameters are categorized into three categories: demographic parameters, obtained from the World Bank ^9^ and the National Institute of Statistics in Tunisia ^10^, biological parameters related to HPV transmission and progression to cervical cancer, and social parameters related to sexual behavior. The latter were obtained from a survey conducted by the Tawhida Ben Cheikh Association on a stratified cluster sample of unmarried Tunisians aged 18 to 30^22^. The details about HPVsim parametrization and a table with parameter and their references is provided in appendix .1.

Unknown model parameter values were evaluate by model calibration using data on HPV genotype prevalence in Tunisia in 2012 national survey^3^ and yearly cancer cases from 2020^23^ and 2022^4^. The method used was Adam^24^. Details on the calibration methodology is provided in appendix .2

We used evidence to estimate the level of protection of Bivalent HPV vaccine against the HPV genotypes that are prevalent in Tunisia^25,26^ (genotypes 16, 18, 31, 35, 39, 52, 56, 58 and 58). This information was used to update the cross-protection matrix of the vaccine in the HPVsim model. The appendix .4 provides the updated cross-protection matrix.

### Intervention scenarios

In January 2024, the Ministry of Health in Tunisia announced plans to introduce the HPV vaccine into the national school vaccination program starting in 2025. The vaccine will be administered to girls in their 6th year of primary school^11^ (11-12 years old). With a vaccination coverage rate of approximately 97% in school settings and a school enrollment rate of 92%, the program is expected to provide widespread protection against HPV. A prior cost-effectiveness analysis on vaccine selection suggested that the bivalent vaccine, priced under 10.5 USD, is a cost-efficient choice^5^.

Currently, cervical cancer screening in Tunisia is estimated to cover 17% of women aged 35 to 60 which translates to approximately a yearly screening coverage rate of 1%, using Pap-smear cytology. Women who test positive for high-risk HPV types are referred for further evaluation through colposcopy and biopsy, where precancerous lesions can be identified and then treated. A detailed description of the screening and treatment process is provided in appendix .3. The low screening coverage is mainly due to the limited capacity of laboratories to process Pap-smear tests. Consequently, there is growing interest in increasing coverage by introducing the HPV DNA test, which can be automated and processed without manual intervention.

For this study, we aim to evaluate the costs and benefits of the following four scenarios: maintaining the current screening strategy coupled with vaccination (Scenario 1); introducing the HPV DNA test at a frequency of one screening during a lifetime between the ages of 35 and 40, coupled with vaccination (Scenario 2); introducing the HPV DNA test at a frequency of two screenings during a lifetime spaced 5 years apart between the ages of 35 and 45, coupled with vaccination (Scenario 3); and finally, introducing the HPV DNA test at a frequency of every 5 years for women aged 35 to 60, coupled with vaccination (Scenario 4). For every scenario that includes the introduction of the HPV DNA test, we assumed a gradual rollout beginning in 2030, with the desired final coverage rate being reached in 2035, thereby fully replacing the Pap-smear test. We tested different targeted coverage rates: 15%, 25%, 33%, 50%, and 70%.

These scenarios will be compared to a Baseline scenario defined as follows: maintaining the current screening strategy with a 1% screening rate per year for women aged 35 to 60 years, without introducing any modifications to the screening methodology or rate, and without introducing vaccination.

For all scenarios, including the Baseline scenario, we assumed that 90% of those who test positive for HPV would follow up with colposcopy and biopsy, and 90% of those diagnosed with precancerous lesions would receive appropriate treatment. This assumption includes a 10% drop-off at each stage to approximately represent those who do not continue treatment. We also suppose that vaccination will be integrated into the existing school vaccination program using the bivalent vaccine, targeting girls aged 11 to 12, at an estimated coverage rate of 90%, starting in 2025.

### Costs

All costs are of 2024 adjusted with a 3% discount rate applied for future years. The bivalent vaccine was priced at 10.25USD per dose^5^. This cost includes delivery and implementation costs (necessary infrastructure, logistics, personnel, refrigeration units, etc …). The treatment cost for cervical cancer was estimated at 2463.00USD per case^27^, the cost has been applied to all cancer cases, assuming that all cancers are being treated. Additionally, the costs for pap-smear test, colposcopy and pre-cancer treatments, including excision and ablation, were based on the official CNAM Hospital Price Template^12^ at 6.50USD, 6.80USD, 18.00USD, and 18.00USD, respectively. CNAM is the National Health Insurance Fund, established to manage health insurance systems and provide coverage for medical expenses for Tunisian citizens. Finally the HPV DNA test was estimated at 29.00USD per test based on expert consultation. All costs are summarized in Table 3 in appendix .2.

### Outcomes

The simulations were run from 1990 to 2090 to compute the full effects of transmission dynamics and intervention outcomes, with each scenario being simulated 10 times for each coverage rate. The primary outcomes of HPVsim include the yearly number of HPV cases, cancer cases, vaccine doses administered, tests performed, colposcopies conducted, and pre-cancer treatments administered. These outcomes were collected from 2025 to 2090. The yearly cancer cases and HPV infections from HPVsim were fitted using a 5th degree polynomial for smoothing; this was applied across all 10 replicates for each coverage rate in each scenario. Costs were calculated by multiplying the yearly HPVsim outcomes of each medical intervention by its corresponding unit price, assuming that all cervical cancer cases would receive treatment. This was also applied across all 10 replicates for each coverage rate in each scenario.

We performed random sampling on the data for averted HPV infections (yearly and cumulative), averted cancer cases (yearly and cumulative), increased costs (yearly and cumulative), and cost-effectiveness as follows: We selected a group of 3 replicates from the 10 replicates of a given coverage rate in each scenario and then averaged the results. This process was repeated 60 times for each coverage rate in every scenario. To determine the averted cancer cases of each replicate, we calculated the reduction in cancer cases compared to the average of the baseline scenario. We assessed the associated cost increase of each replicate by comparing the cost to the average of the baseline scenario. We defined cost-effectiveness using the following criteria: a scenario is considered highly cost-effective if the increased cost per cancer case averted is less than one times the average per capita Tunisian gross domestic product (GDP); cost-effective if it is between one and three times the average GDP; and not cost-effective if it exceeds three times the average GDP. The per capita Tunisian GDP is estimated to be 3895USD in 2023^28^, discounted by 3% every year from 2025 to 2090 and then the average was calculated.

## Supporting information

Model Parameter 1

Model Parameter 2

Model Parameter 3

Model parameter 4

## Data Availability

All data produced in the present work are contained in the manuscript.

## Acknowledgements

This work was supported, by Bill Melinda Gates Foundation [INV-059607] through AMAX (African Modeling and Analytics Academy for Women) project and by Vaccine Impact Modelling Consortium (VIMC). Under the grant conditions of the Foundation, a Creative Commons Attribution 4.0 Generic License has already been assigned to the Author Accepted Manuscript version that might arise from this submission. At the time of analysis, the VIMC was jointly funded by Gavi, the Vaccine Alliance and the Bill Melinda Gates Foundation (grant numbers INV-034281 and INV-009125/OPP1157270).

## Author contributions statement

AL conducted the simulations, performed result analysis, as well as contributed to article drafting. BB assisted in data collection and contributed to writing the discussion draft. AG, EE, RS and SRR provided recommendations and suggestions for conceiving the simulations. RS and SRR reviewed the first draft of the article. OL helped with data analysis. AK, HA and SBM provided supervision, with SBM also contributing to the methodology and article drafting.

## Additional information

### Genotype Identification

In the context of our simulation, it is essential to accurately identify and model the different genotypes of the HPV virus and their distribution within the Tunisian population. We based our study on a survey conducted in Tunisia in 2013, aimed at determining the distribution of HPV genotypes in the country. The genotypes identified in this survey included the following: HPV-6, 11, 16, 18, 31, 35, 39, 40, 42, 43, 44, 52, 53, 54, 56, 58, 59, 62, 66, 68, 70, 75, 81, 84, 89. It is important to note that these genotypes include both high-risk types, i.e., those that can cause cancer, and low-risk types, which do not cause cancer. To better understand their potential impact on public health, it is crucial to know the used classification to classify substances according to their carcinogenic potential. Here is an explanation of the different classes :

- Group 1 Carcinogens: This category includes substances that are recognized as carcinogenic to humans. The HPV genotypes included in this category are those most closely associated with the development of cervical cancer
- Group 2A Carcinogens: Substances in this category are probably carcinogenic to humans. There is sufficient evidence of their carcinogenicity in animals and limited evidence in humans
- Group 2B Carcinogens: Substances in this category are possibly carcinogenic to humans. There is limited evidence of their carcinogenicity in humans and insufficient evidence in animals
- Phylogenetic Analogies with Carcinogenic Types: This classification includes HPV types that, although not directly proven to be carcinogenic, have phylogenetic similarities with known carcinogenic types
- Non-Carcinogenic: This category includes HPV types that have not been associated with a cancer risk

Applying this classification, we first eliminated the genotypes known to be non-carcinogenic. Then, using the classification presented in table 1, derived from an in-depth study of HPV genotypes responsible for cancer, we refined our selection. Ultimately, we retained the following carcinogenic genotypes for our simulation: HPV-16, 18, 31, 35, 39, 52, 56, 58, 59. The other genotypes were not completely excluded from the model but were grouped into a cluster representing low-risk genotypes

**Table 1.** Classification of HPV genotypes using the classification found in^29^.

| **Carcinogenic Potential Class** | **Genotypes** |
| --- | --- |
| Group 1 | 16, 18, 31, 33, 35, 39, 45, 51, 52, 56, 58, 59 |
| Group 2A | 68 |
| Group 2B | 26, 53, 66, 67, 70, 73, 82 |
| Phylogenetic Analogies | 30, 34, 69, 85, 97 |

The main challenge we faced was that the HPVsim model initially only allowed the selection of HPV-16 and HPV-18 genotypes. Therefore, we had to modify the source code of the HPVsim model to include the newly identified genotypes while ensuring no errors or bugs were introduced. With these modifications, our model can now more accurately represent the diversity of HPV genotypes present in Tunisia. This improves the precision of our simulations and provides more reliable results regarding the transmission and evolution of the virus, which is crucial for evaluating the potential impact of the vaccination and screening strategies we aim to implement.

#### .1. Model Parameterization

##### .1.1 Choice of Simulation Parameters

Multi-agent models and graph-based models are used to represent complex systems where the interactions between agents or nodes are dynamic and often influenced by random processes. Due to this random component, these simulations are inherently stochastic, meaning that the results can vary from one execution to another. To mitigate uncertainty, it is common to do large number of replicates or/and increase the number of agents. This allows for capturing the inherent variability and providing a more accurate estimation of the average results. For this we conducted an analysis where we tested different agent populations 10000, 50000, 100000 and different numbers of replicates: 1, 2, 3, 5, 10, 15, 20. We run HPVsim with the desired number of replicates 10 times, and at each run we calculate the mean and standard deviation of cancer cases resulting from running that number of replicates. We define the centroid as the average value we get from the 10 runs. This process was repeated for every agent population size we set to test.

The analysis on the mean of cancer cases outcomes showed that for a population of 10000 agents the position of the centroid of the scatter plot becomes stable starting from 5 replicates, although the dispersion of the scatter plot becomes significantly reduced at 15 replicates (see Figure 6a). For a population of 50000 agents, the centroid stabilizes at 5 replicates, and the dispersion of the scatter plot becomes significantly less dispersed also at 5 replicates (see Figure 6b). Regarding a population of 100000 agents, the centroid is stable even at one replicate, but dispersion of the scatter plots reduces only after 5 replicates (see Figure 6c). To summarize, to ensure mean stability and minimize disparity (i.e., standard deviation of means), it is recommended to perform 15 replicates for a population of 10000 agents, 5 replicates for population of 50000 agents and 5 replicates for population of 100000 agents.

The analysis on the standard deviation of cancer cases outcomes showed that For a population of 10000 agents, centroid stabilizes at 10 replicates, but the scatter plot remains dispersed even with 20 replicates (see Figure 7a). For a population of 50000 agents, the centroid stabilizes after 10 replicates, and the dispersion is also reduced at 10 replicates (see Figure 7a). For a population of 100000 agents, the standard deviation is stable after 10 replicates however the dispersion is reduced after 5 replicates only (see Figure 7a). To ensure standard deviation stability and minimize disparity (i.e., standard deviation of standard deviations), it is recommended to perform 20 replicates for a population of 10000 agents and 10 replicates for populations of 50000 or 100000 agents.

As a result of all the above, we have identified three options in order to ensure statistically stable results: 30 replicates for a population of 10000 agents, or 10 replicates for populations of 50000 or 100000 agents. Therefore to choose between the valid options it’s crucial to consider runtime. Figure 8 shows the evolution of running time as the number of replicates increases for each agent population size. It is clear that 10 replicates for 50000 agents take much less time than 10 replicates for 100000 agents.Although 20 replicates for 10000 agents take almost as much time as 10 replicates for 50,000 agents, we opted for 10 replicates with 50,000 agents, as this configuration ensures a better statistical stability.

##### .1.2 Calculation of Age of Sexual Debut

The age of sexual debut was derived from data collected in the 2022 survey conducted by the Tawhida Ben Cheikh Group, focusing on unmarried Tunisians aged 18 to 30. Initially, we calculated the average age of sexual debut reported by survey participants, which included both men and women. To adjust for broader population dynamics, particularly considering marriage norms, we weighted these averages using marriage age data provided by the National Institute of Statistics for young Tunisians (aged 18 to 30). This adjustment aimed to more accurately reflect when individuals typically initiate sexual activity in relation to marriage. As a result, the adjusted average age of sexual debut was determined as 23 years for women and 22 years for men. These calculations form the foundation for modeling sexual behavior dynamics in our study, acknowledging the influence of cultural norms and survey limitations that may affect the precision of these estimates.

#### .2. Calibration

The genotype distribution data we have is from the year 2013^3^. Since our model begins the simulation in 1990, we cannot simply input this data directly as the initial distribution. Given the dynamic nature of the model, the genotype distribution evolves over time based on the interactions and parameters of the model. Therefore, it is necessary to define a different initial distribution that, after simulation, will correspond to the observed distribution in 2013. To achieve this, we manually adjusted the initial distribution of genotypes. This process involved numerous iterations and fine adjustments until the resulting distribution in 2013 satisfactorily matched the observed data. The results of this calibration are presented in Figure 9.

Calibration was performed on data of yearly cancer cases in Tunisia from 2020^23^ and 2022^4^. To calibrate our model, we adjusted several parameters whose values were uncertain due to a lack of specific data. These parameters include: transmission probability (beta), relative transmissibility from males to females in penetrative sex (transm2f), relative transmissibility from females to males in penetrative sex (transf2m), male infidelity rate (m_cross_layer) and female infidelity rate (f_cross_layer). HPVsim includes a calibration tool based on random sampling of values and uses the mean absolute error as a fitting metric. However, this default algorithm has limitations, including non-guaranteed convergence and very long computation times. To overcome these limitations, we developed our own calibration function using the optimized stochastic gradient algorithm ADAM. This algorithm also uses mean absolute error as the performance metric. Thanks to this approach, we were able to achieve and efficient convergence. The results of this calibration are illustrated in Figure 10, which shows that the forecasts generated by HPVsim after calibration are within a 10% margin of error compared to national forecasts for new cancer cases, and the curve has almost the same slope as the curve of cases from the national predictions. This accuracy confirms the effectiveness of our calibration method and ensures that the model reliably reflects current and future cancer case trends in Tunisia.

#### .3. Current HPV Screening and Treatment Process

HPV screening in Tunisia follows a well-defined clinical process. Screening begins with a Pap test, also known as a cervical smear test. This test involves collecting cells from the cervix and examining them under a microscope to detect abnormalities. The main goal of the Pap test is to early identify cellular changes that could develop into cervical cancer, thus allowing for quick and preventive intervention. When Pap smear test results reveal cellular abnormalities, the patient is referred for a colposcopy with a biopsy. A colposcopy is a procedure where a colposcope is used to closely examine the cervix for suspicious areas. A biopsy, which involves taking a small tissue sample, is performed for a more detailed histological analysis. The results of the colposcopy and biopsy help determine the type of lesions present:

- LSIL (Low-grade Squamous Intraepithelial Lesion): LSIL are mild cellular changes, often caused by a transient HPV infection. Although these lesions have a low potential to become cancerous, they require monitoring. The typical treatment for LSIL is ablation, a procedure that removes or destroys the abnormal tissue using a laser.
- HSIL (High-grade Squamous Intraepithelial Lesion): HSIL indicates more severe cellular changes that have a higher risk of progressing to cervical cancer. HSIL treatment involves a conization procedure, which removes a cone-shaped portion of the cervix containing the abnormal cells.
- Cancer: If the biopsy reveals the presence of cancer cells, the patient is immediately referred for appropriate cancer treatment, which may include a combination of surgery, radiotherapy, and chemotherapy, depending on the cancer’s stage and location.

This systematic process helps identify and treat cellular abnormalities at an early stage, thus reducing the progression to more severe forms of the disease.

#### .4. Vaccine Cross-protection

This appendix provides the updated cross-protection values (Table 2) of Bivalent vaccine against different HPV genotypes using one dose. We assumed a 100% efficacy for the targeted genotypes by a given vaccine, although this may be overestimated, we believe it won’t affect the outcomes of the study as the vaccination intervention will be the same across the tested screening strategies. We also assumed a 0% efficacy in case we didn’t find any evidence of protection.

**Table 2.**
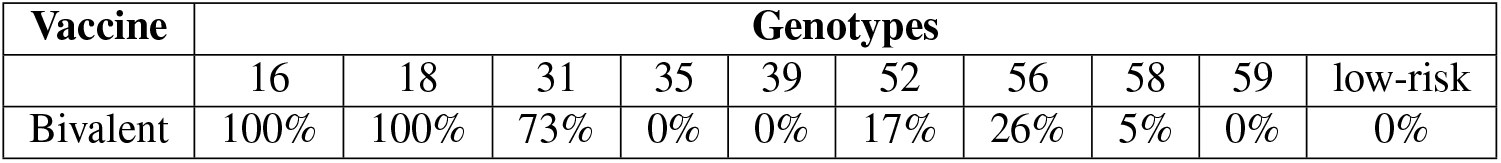
Cross-protection of different HPV vaccines against various genotypes.

| Vaccine | Genotypes |  |  |  |  |  |  |  |  |  |
| --- | --- | --- | --- | --- | --- | --- | --- | --- | --- | --- |
|  | 16 | 18 | 31 | 35 | 39 | 52 | 56 | 58 | 59 | low-risk |
| Bivalent | 100% | 100% | 73% | 0% | 0% | 17% | 26% | 5% | 0% | 0% |

#### .5. Interventions Costs

This appendix offers a detailed summary of unit costs (Table 3). It outlines the expenses associated with various healthcare interventions for HPV infections and cervical cancer, reflecting 2024 pricing and applying a 3% discount rate for future years.

**Table 3.** Summary of Healthcare Intervention Unitary Costs. CNAM referred to the Tunisian National Health Insurance Fund.

| Healthcare Intervention | Unitary Cost (USD) in 2024 | Source |
| --- | --- | --- |
| Bivalent vaccine (per dose) | 10.25 | Source<sup>5</sup> |
| Cervical cancer treatment (per case) | 2463.00 | Source<sup>27</sup> |
| Pap smear test | 6.50 | CNAM Hospital Price Template |
| Colposcopy | 6.80 | CNAM Hospital Price Template |
| Pre-cancer treatments (excision) | 18.00 | CNAM Hospital Price Template |
| Pre-cancer treatments (ablation) | 18.00 | CNAM Hospital Price Template |
| HPV DNA test | 29.00 | Expert consultation |

#### .6. Model Parameters

HPVsim includes various categories of parameters, including simulation parameters, demographic parameters, biological parameters, and behavioral parameters. In this appendix, we present the parameters customized for our study, detailed in Tables 4, 5. Parameters not listed in this appendix were set to their default values.

**Table 4.**
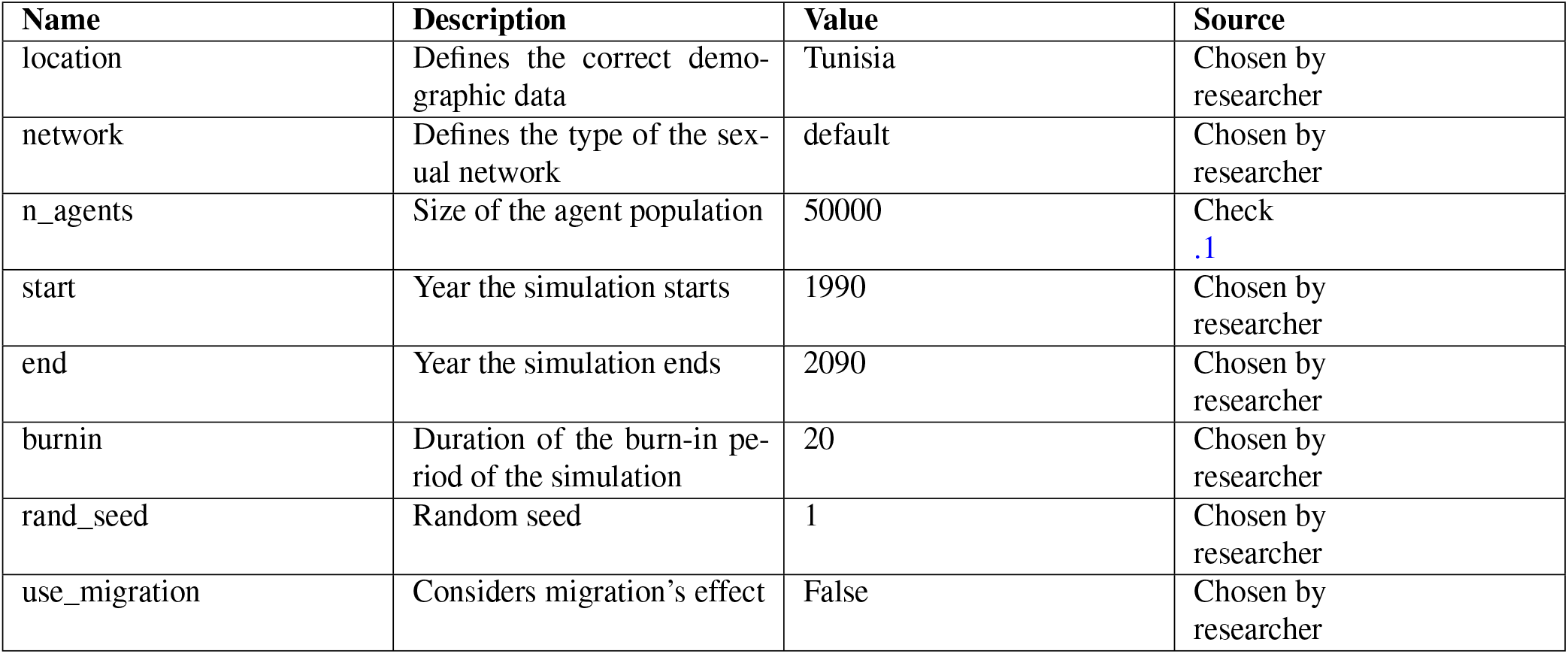
HPVsim parameters and values used for the simulations 1/2.

| Name | Description | Value | Source |
| --- | --- | --- | --- |
| location | Defines the correct demographic data | Tunisia | Chosen by researcher |
| network | Defines the type of the sexual network | default | Chosen by researcher |
| n_agents | Size of the agent population | 50000 | Check .1 |
| start | Year the simulation starts | 1990 | Chosen by researcher |
| end | Year the simulation ends | 2090 | Chosen by researcher |
| burnin | Duration of the burn-in period of the simulation | 20 | Chosen by researcher |
| rand_seed | Random seed | 1 | Chosen by researcher |
| use_migration | Considers migration's effect | False | Chosen by researcher |

**Table 5.** HPVsim parameters and values used for the simulations 2/2.

| Name | Description | Value | Source |
| --- | --- | --- | --- |
| init_hpv_prev | Initial age distribution of HPV infections | **age_brackets:** [30, 40, 50, 150]<br>**m:** [0.124, 0.07, 0.064, 0.081]<br>**f:** [0.124, 0.07, 0.064, 0.081] | Source<sup>3</sup> |
| init_hpv_dist | Initial HPV genotypes distribution | **hpv31:** 0.019<br>**hpv16:** 0.011<br>**hpv59:** 0.0125<br>**hpv52:** 0.012<br>**hpv18:** 0.011<br>**hpv35:** 0.0055<br>**hpv58:** 0.0065<br>**hpv56:** 0.0045<br>**hpv39:** 0.0045<br>**lr:** 0.9135 | Check .2 |
| debut | Age of sexual debut | **f:** ('normal', par1=23, par2=3)<br>**m:** ('normal', par1=22, par2=4) | Check appendix .1 |
| Name | Description | Value | Source |
| beta | Transmission coefficient | 0.13 | Calibration value, check .2 |
| transm2f | Male-to-female transmission multiplier | 3 | Calibration value, check .2 |
| transf2m | Female-to-male transmission multiplier | 1 | Calibration value, check .2 |
| eff_condoms | Condom efficiency | 0.7 | Source<sup>30</sup> |
| condoms | Condom usage rate for different types of relationships | **m:** 0.01<br>**c:** 0.18 | Source<sup>22</sup> |
| m_cross_layer | Male cross-layer mixing parameter | 0.25 | Calibration value, check appendix .2 |
| f_cross_layer | Female cross-layer mixing parameter | 0.1 | Calibration value, check appendix .2 |
| hpv_control_prob | Probability of HPV control | 0 | Chosen by researcher |
| Name | Description | Value | Source |
| dur_pship | Duration of relationships | **m:** ('lognormal', par1=20, par2=26)<br>**c:** ('lognormal', par1=1, par2=2) | Chosen by researcher |
| layer_probs | Share of females and males of each age who are actively seeking relationship if under-partnered | See Excel files: layer_probs_m.xlsx and layer_probs_c.xlsx | Source<sup>31</sup> |
| mixing | Age mixing matrices for marital and casual relationships | See Excel files: mixing_m.xlsx and mixing_c.xlsx | National Institute of Statistics |

#### Footnotes

1 https://www.afro.who.int/health-topics/cervical-cancer

2 https://www.who.int/europe/news/item/11-09-2021-who-recommends-dna-testing-as-a-first-choice-screening-method-for-cervical-cancer-prevention

3 https://supply.unicef.org/s0001704.html

4 https://www.hologic.com/sites/default/files/MISC-06024-001_001_01.pdf

5 https://www.fishersci.com/shop/products/onclarity-hpv-test/B441990

6 https://www.tap.info.tn/fr/Portail-SociÃl’tÃl’/17851371-le-vaccin-contre-le

7 https://www.who.int/europe/news/item/11-09-2021-who-recommends-dna-testing-as-a-first-choice-screening-method-for-cervical-cancer-prevention

8 https://applications.emro.who.int/docs/Country_profile_2013_EN_15402.pdf

9 https://data.worldbank.org/country/tunisia

10 https://www.ins.tn/taxonomy/term/113

11 https://www.tap.info.tn/fr/Portail-SociÃl’tÃl’/17851371-le-vaccin-contre-le

12 https://www.cnam.nat.tn/doc/upload/RBactes.pdf

